# Deciphering rifampicin resistance among tuberculosis patients who have trace results on Xpert MTB/RIF ultra assay

**DOI:** 10.1101/2025.06.13.25329610

**Authors:** Nelissa Juliet Nantumbwe, Derrick Semugenze, Francis Okello, James Sserubiri, Kevin Komakech, Benon Asiimwe, Willy Ssengooba

## Abstract

**Introduction:** The *Mycobacterium. tuberculosis* (MTB) detected trace, rifampicin (RIF) resistance indeterminate category of the Xpert MTB/RIF Ultra assay results is usually, non-actionable and requires retesting the samples. We aimed to decipher RIF resistance among tuberculosis patients who have trace and indeterminate results.

**Methods:** Four hundred and three (403) MTB detected trace, RIF resistance indeterminate results, which were obtained in Mycobacteriology (BSL-3) and Molecular Diagnostic laboratories, College of Health Sciences, Makerere University from August 2018 to June 2023, having culture results were identified from the laboratory database. Isolates of those that turned out culture positive were retrieved and sub-cultured in liquid media to perform phenotypic first line Drug Susceptibility tests, first line Line-Probe assays (LPA) and repeat GeneXpert ultra.

**Results:** A total of 31/403 (7.7%) culture positive isolates were identified from the database of which 77.42% (24/31) were positive for *Mycobacterium tuberculosis* complex (MTBc). Nineteen (19) out of the identified 24 MTBc were successfully retrieved, cultured and resistance testing performed. Phenotypic Drug susceptibility testing and repeated GeneXpert did not identify any resistance. Only one mutation *inhA* MUT1 related to isoniazid (INH) resistance was identified using MTBDRplus assay.

**Conclusion:** In this study, we did not identify any missed rifampicin resistance among MTBc culture positive samples that were initially Xpert ultra-trace and rifampicin resistance indeterminate. More studies with bigger sample sizes especially in high MDR-TB settings are required.

## Introduction

Tuberculosis is a communicable disease which accounts for the highest mortality from any infectious diseases worldwide surpassing Covid. In 2023, an estimated 10.8 million people developed Tuberculosis with an incidence rate of 134 per 100,000 population and 189,693 were people diagnosed with drug-resistant Tuberculosis(1). In Uganda, there were 86,469 people newly diagnosed with Tuberculosis with an incidence rate of 198 per 100 000 population with 660 people being diagnosed with Drug resistant TB. The Xpert MTB/RIF Ultra assay (Ultra: Cepheid, Sunnyvale, CA, USA) was endorsed by WHO in 2017 an advanced version of the Xpert MTB/RIF endorsed in 2012, which detects *Mycobacterium tuberculosis* using probes that target multicopy *1S6110* and *IS1081*. Detection of these two targets and optimized polymerase chain reaction cycling increases the sensitivity of Ultra for detection of MTBc(2). This assay also rapidly determines rifampicin resistance by detecting mutations within the 81 base pair rifampicin-resistance determining region (RRDR) within the *rpoB* gene of MTBc(3). There is detection of *IS6110* and *IS1081* in samples with trace calls however, the signal for rifampicin resistance determination is insufficient(5) which could be due to low bacillary load in the tested samples(4) or due to dead bacilli(5).

Just like any other diagnostic test, increasing the sensitivity of the Ultra assay and also the introduction of the trace category led to decreased specificity ((6)(3). Interpretation and application of the trace results in clinical practice is still challenging(5). According to WHO, the trace results are considered bacteriologically confirmed tuberculosis for people living with HIV (PLHIV), children who are being evaluated for pulmonary TB, individuals being considered for extra pulmonary tuberculosis, and non-HIV individuals who have never had anti-TB treatment in the last 5 years(5). It is also worth noting that WHO does not recommend repeating these results (7). However much the clinicians can begin treatment for the specified categories, they do so blindly without knowing the rifampicin resistance status since it is indeterminate. There is documented evidence of discordant rifampicin resistance results between the Ultra assay and MTBDRplus (Hain Lifescience GmbH, Nehren, Germany) on samples that were initially MTB detected trace and RIF resistance indeterminate by the Ultra assay (8). Employing both methods therefore. may capture rifampicin resistance that may be missed by one method when determining the proportion of rifampicin resistance among individuals who initially had trace calls and rifampicin resistance indeterminate.

## Methods

### Patients and settings

This study was approved by the Makerere University, School of Biomedical Sciences Research and ethics Committee (SBS-2022-244) on 29^th^ March 2023. This was a cross-sectional laboratory-based study conducted at the College of American Pathologists (CAP: ISO15189) Accredited Mycobacteriology (BSL-3) Laboratory, Department of Medical Microbiology Makerere University College of Health Sciences. Kampala, Uganda. We searched the Mycobacteriology BSL-3 Laboratory’s database on 03rd April 2023 for MTB detected trace RIF resistance indeterminate results obtained between August 2018 (since obtaining the GeneXpert Ultra system) to June 2023. Four hundred and three (403) results were retrieved. From these, we purposively sampled out all culture positive results and retrieved their corresponding culture isolates. The experiments were ran until 30^th^ November 2023 and authors had no access to information that could identify individual participants during or after data collection.

### Isolate sub-culturing

Isolates were retrieved from the −80°C freezer, thawed, and sub-cultured. Briefly 0.5mls of the isolates were inoculated in the Mycobacterial growth Indicator Tubes (MGIT) broth with a growth supplement and incubated at 37°C in the BACTEC™ MGIT™960 System (BD Franklin Lakes, USA) from which only pure positive subcultures were used for Streptomycin, Isoniazid, Rifampicin, Ethambutol (SIRE) phenotypic drug susceptibility testing in MGIT machines, tested for Isoniazid and Rifampicin using MTBDRplus assay and repeat Ultra assay.

### Phenotypic Drug Susceptibility Testing (pDST) using the MGIT system

Drug vials of Isoniazid (INH), Rifampicin (RIF), Ethambutol (ETH) and streptomycin (STR) were reconstituted each with 4 ml of sterile deionized (DI) water, and mixed well. Eight hundred microliters (800 µl) of an antimicrobial mixture of polymyxin B, amphotericin B, nalidixic acid, trimethoprim, and azlocillin (PANTA) (Becton Dickinson Diagnostic Systems, Sparks, MD) was added into five MGIT medium (7-ml bar-coded MGIT tubes) tubes containing a growth supplement. One hundred microliters (100µl) of each drug for each sample was added into respective labelled MGIT medium (7-ml bar-coded MGIT tubes) tubes. A 0.5 McFarland was prepared for each sample to be used for carrying out pDST. A drug free “ growth control tube” that has PANTA was inoculated with each isolate with a prepared McFarland diluted 1:100. For each sample, 500µl of the McFarland sample was added into respective tubes labelled STR, INH, RIF and ETH to make final concentrations of 1.0 µg/ml, 1.0 µg/ml, 0.5 µg/ml and 5.0 µg/ml respectively for these drugs. These inoculated tubes were put on the AST carrier, scanned into the BACTEC™ MGIT™960 System (BD Franklin Lakes, USA), and incubated at 37°C. The interpretation was done automatically by the machine when the growth units were below 100 GU.

### Drug Susceptibility Testing for Isoniazid and Rifampicin using MTBDRplus assay

This was performed directly on the sub subcultured samples, which were MTBc positive non non-contaminated per the manufacturer’s guidelines as described in the GenoType MTBDR*plus* VER 2.0 kit (10) and as detailed by Sharma (9). Briefly, 45 μL of master mix per DNA template was prepared in the contamination-free master mix room, and 5 μL of DNA template was added in the template addition room. Amplification was carried out using a two-step multiplex PCR that was composed of biotinylated primers for reverse hybridization with specific probes. Amplification and hybridization were performed for MTBDRplus V2.0 (Hain Lifescience GmbH, Nehren, Germany). Each strip was evaluated for validity by the presence of control bands. A positive result for TB was indicated by the presence of a band against the MTBc control line on the strip

### Repeat Xpert MTB/RIF Ultra assay

This was done on all sub-cultured results that were MTBc positive, Non-Tuberculous Mycobacterium (NTM) and contaminated samples as per the Standard Operating Procedures (SOP) of the lab and the manufactures package insert (10).

### Statistical analysis

All trace call results were exported from the GeneXpert database into Microsoft Excel and the respective demographics, phenotypic DST, MTBDRplus V2.0 and repeat Xpert MTB/RIF Ultra assay results were updated. All statistical analysis was conducted from Stata software, version 15. Percentages, means, medians, and Interquartile ranges were calculated for the different demographics and presented. P-values and confidence intervals were not obtained due to the nature of our findings.

## Results

### Baseline and clinical characteristics of the participants

Four hundred and three (403) MTB detected trace; RIF resistance indeterminate results were obtained from the laboratory’s database. A total of 31/403 (7.7%) culture positive isolates were identified, of which 77.42% (24/31) were positive for *Mycobacterium tuberculosis* complex (MTBc). Nineteen (19) out of the identified 24 MTBc were successfully retrieved, cultured and resistance testing performed. As shown in Table 1 below, the median age (years) of the study group was 32 (IQR 25, 40). Males accounted for 56.82% (229/403). Most samples retrieved were screening samples (35.74%), and the least were follow-up samples, which ranged from Week 0 to Week 50. Samples that were tested and resulted into trace calls included; sputum (87.59%), stool (4.71%), nasal swabs (2.73%), gastric aspirate (1.74%), cerebrospinal fluid (0.74%), isolates (0.50%), nasal pharyngeal aspirate and pleural fluid each accounted for 0.25%.

**Table 1:** Baseline and clinical characteristics of the participants.

| Characteristics | Category | Frequency (403) | Percentage (%) |
| --- | --- | --- | --- |
| **Gender** | Female | 142 | 35.24 |
|  | Male | 229 | 56.82 |
|  | Not provided | 32 | 7.94 |
| **Visit interval** | Baseline | 89 | 22.08 |
|  | Follow-up | 37 | 9.18 |
|  | Not indicated | 133 | 33.0 |
|  | Screening | 144 | 35.73 |
| **Sample type** | Cerebrospinal Fluid | 3 | 0.74 |
|  | Gastric Aspirate | 7 | 1.74 |
|  | Isolates | 5 | 1.24 |
|  | Nasopharyngeal Aspirate | 1 | 0.25 |
|  | Nasal Swab | 11 | 2.73 |
|  | Pleural Fluid | 1 | 0.25 |
|  | Sputum | 353 | 87.59 |
|  | Stool | 19 | 4.71 |
|  | Not documented | 3 | 0.74 |

### Subculture Results

Of the 31 culture positive isolates in the database with trace call, 24 (77.42%) were confirmed MTBc positive while 7 (22.58%) were NTM’s. Majority of the MTBc positive were sputum samples (87.5%). The analysis of 24 confirmed MTBc isolates provided a snapshot of the sample characteristics. The median age for the samples was 38 years (IQR 29, 44) with the male gender being 14 (66.67%). Out of the 24 MTBc positive isolates, only 19 were accessible in −80°C. These were thawed and MGIT sub cultured and hence subsequent experiments were done on these.

### Rifampicin resistance-conferring mutation determined by MTBDRplus assay

We did not find any rifampicin resistance-conferring mutation detected on all the 19 direct MTBc positive isolates. We however, found only one mutation *inhA* MUT1 associated with isoniazid resistance.

### Rifampicin resistance detected by the repeat Xpert MTB/RIF Ultra assay

19 isolates were subjected to a repeat GeneXpert Ultra, and as shown in Figure 1 below, the biggest percentage (42.11%) of the results obtained were MTB detected medium; RIF resistance not detected and the lowest was MTB not detected (5.26%). No single repeat sample was MTB detected trace; RIF resistance Indeterminate.

**Figure 1:**
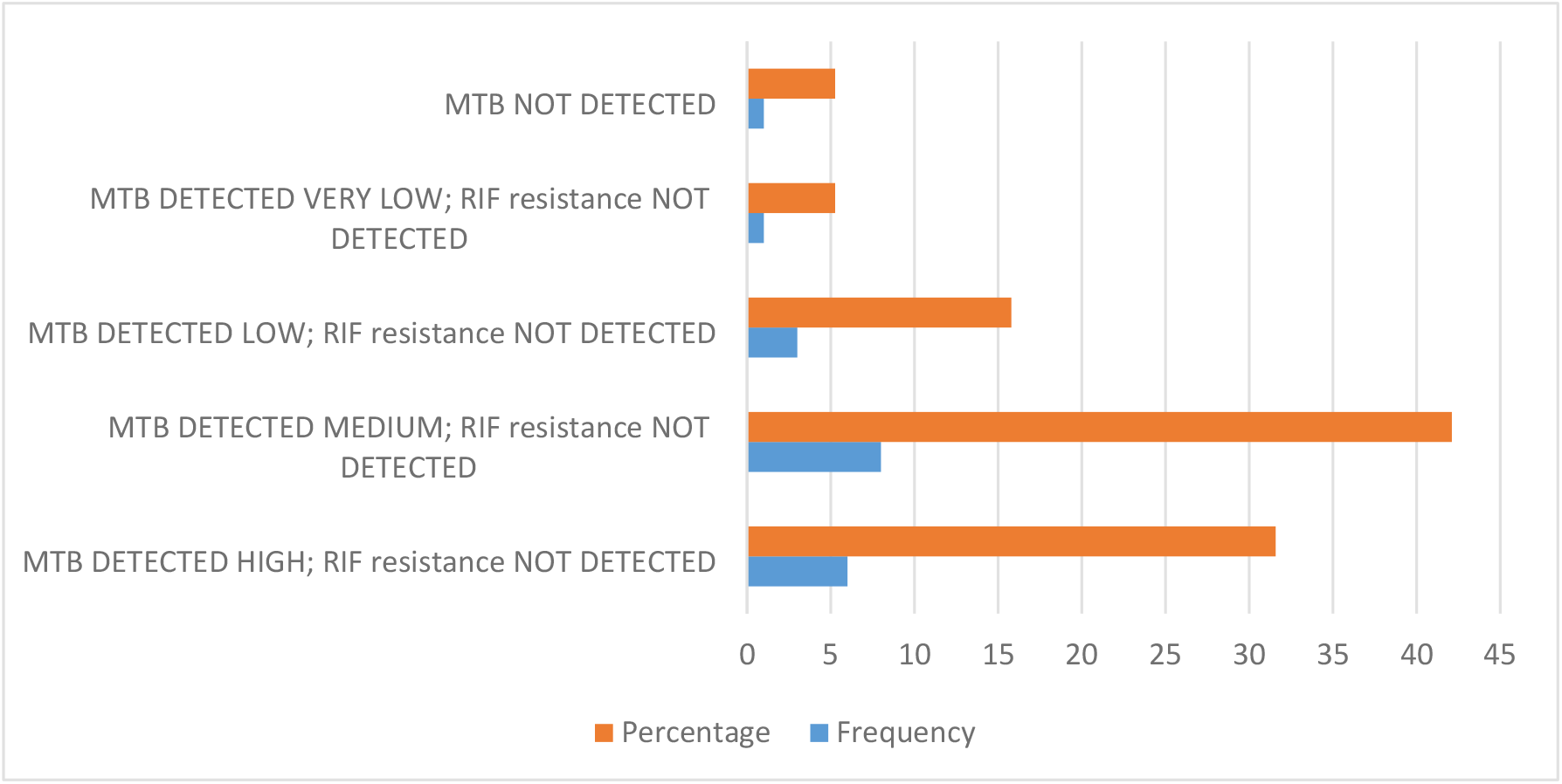
Repeat Xpert MTB/RIF Ultra assay results done on sub-cultured isolates

### Rifampicin resistance determined by pDST

We did not find any rifampicin resistance that was determined by carrying out pDST on all the tested 19 MTBc positive isolates. There was resistance detected to Isoniazid from 2 culture positive isolates

## Discussion

The present study aimed to decipher rifampicin resistance among tuberculosis patients who have trace call results on Xpert MTB/RIF ultra-assay. Our study revealed no missed rifampicin resistance among patients with the MTBDRplus assay, repeat ultra and pDST among isolates from participants who initially had ultra-trace and rifampicin indeterminate results. These same results are in agreement with a study done by Huang *et al* (4)where there was concordance between Ultra and Line Probe Assay. In this study, Ultra identified rifampicin resistance more accurately than Xpert, when corroborated against other methods such as broth microdilution and line probe assay. Our study however, showed that there was no false rifampicin resistance when the xpert MTB/RIF ultra-assay was repeated. This was in agreement with a study done by Ngabonziza *et al*(11) who reported 47% of the tested samples to have false rifampicin resistance. The absence of the rifampicin resistance detection or any of its conferring mutations being detected in our tested samples could have been due to Uganda being a low MDR-TB setting where there was a reported 1.2% and 12.1% prevalence of drug-resistant TB among new and retreatment cases respectively during a national survey carried out in 2010 (12) or as an indicator of underlying *rpoB* mutations within the rifampicin resistance determining region (2). More to this, even the number of MTBc positive samples (n=19) that was found among the trace call results in our study was really low to capture some rifampicin resistance, given the reported prevalence of MDR-TB. The small number of trace call results that we obtained from the Xpert MTB/RIF ultra assay database over the reviewed period of approximately 5 years could be due to the strength of the National TB and Leprosy Program (NTLP) moving the TB diagnostic testing using the Xpert MTB/RIF ultra assay to the lower health facilities and hence fewer patients sought these services at central facilities like where our study was carried out. Sputum samples were the predominant tested samples (87.50%) among the confirmed MTBc isolates. This could be due to the assay having been optimized and validated for pulmonary TB diagnosis(3). Among the samples for the diagnosis of pulmonary TB, collection of sputum samples is less invasive. Invasiveness could be such a big factor because next to sputum samples, stool and nasal swabs were among the samples that had high numbers. Only 1.74% and 0.25% of the tested samples for pulmonary TB were gastric aspirates and nasopharyngeal aspirates respectively which could have been due to the required skills for the collection of these samples and the invasiveness of the procedures. Other sample types that were tested were for diagnosis of extrapulmonary tuberculosis (EPTB) which has been known to be paucibacillary in nature(14). These samples included pleural fluid and cerebrospinal fluid. These findings are in agreement with previous studies that highlight EPTB occurring less contagiously than pulmonary TB, and is often ignored during initial diagnosis (13). Related to our study findings, a study done in India (14) who 31.3% of the tested pulmonary samples using MTBDRplus assay had rifampicin indeterminate results however, single double and some triple mutations were found in other regions of the *rpoB* gene upon carrying out DNA sequencing. This would perhaps indicate that there could be some rifampicin conferring mutations outside the hot-spot region that were not captured by the MTBDRplus and Xpert MTB/RIF ultra assays on the samples that we tested. To the best of our knowledge, our study is the first one to investigate this subject in Ugandan settings and hence it is paving way to future studies in this scientific field.

This study was limited by the Trace call sample size; however, trace calls are rare, and this may be one of the largest numbers obtained for a similar analysis. This study was conducted in a single facility which could limit the generalization of the results however, Mycobacteriology BSL-3 laboratory is a facility that receives and analyses samples from different country’ sites so the findings remain valid. Some demographics and sample information were missing in our results which is attributed to the retrospective review of the results in the database. Retrieval and usage of sub-cultured stored isolates could have missed out vulnerable bacilli populations and finally this study was carried out in low MDR-TB settings which might have led to not finding any rifampicin resistance among the tested isolates.

There is a need for future prospective studies with bigger sample size, including settings with higher prevalence of MDR-TB that also involve sequencing isolated *Mycobacterium tuberculosis* to identify mutations in the *rpoB* gene that may not be captured by the Ultra and MTBDRplus assays.

## Data Availability

All relevant data are within the manuscript and its Supporting Information files.

## Acknowledgements

The authors would like to acknowledge the staff of Mycobacteriology (BSL-3) and Molecular Diagnostic laboratories who supported this research by providing all the required data and working with NNJ on the different laboratory procedures.

## Data availability statement

All data generated or analyzed during this study are included in this published article

## Authors’ contributions

NJN, BA, and WS conceived the idea. NJN did the investigation, data curation and formal analysis. All co-authors’ DS, JS, KK and FO were involved in the literature review and methodology. BA and WS did the supervision. Every co-author reviewed the final draft before submission.

